# Early Treatment with Fluvoxamine Among Patients with COVID-19: A Cost-Consequence Model

**DOI:** 10.1101/2021.12.23.21268352

**Authors:** Fergal P. Mills, Gilmar Reis, Kristian Thorlund, Jamie I. Forrest, Christina M. Guo, David R Boulware, Edward J. Mills, for the TOGETHER Investigators

**Affiliations:** InnomarConsulting - Oakville, Canada; Research Division, Cardresearch, Cardiologia Assistencial e de Pesquisa, Brazil; Platform Life Sciences Inc. Vancouver, BC, Canada; Department of Medicine, University of Minnesota, Minneapolis, MN, USA; Department of Health Research Methods, Evidence, and Impact, McMaster University, Hamilton, Ontario, Canada

## Abstract

**Background:** Three randomized trials have been conducted indicating a clinical benefit of early treatment with fluvoxamine versus placebo for adults with symptomatic COVID-19. We assessed the cost-consequences associated with the use of this early treatment in outpatient populations.

**Methods:** Using results from the three completed trials of fluvoxamine vs. placebo for the treatment of COVID-19, we performed a meta-analysis. We conducted a cost-consequence analysis using a decision-model to assess the health system benefits of the avoidance of progression to severe COVID-19. Outcomes of relevance to resource planning decisions in the US and elsewhere, including costs and days of hospitalization avoided, were reported. We constructed a decision-analytic model in the form of a decision tree to evaluate two treatment strategies for high-risk patients with confirmed, symptomatic COVID-19, from the perspective of a third-party payer:(1) treatment with a 10-day course of fluvoxamine (100mg twice daily); (2) current standard-of-care; (3) molnupiravir 5-day course. We used a time horizon of 28 days.

**Results:** Administration of fluvoxamine to symptomatic outpatients with COVID-19 at high-risk of developing progression to severe COVID-19 complications is substantially cost-saving in the US, in the amount of $232 per eligible patient, and saves an average of 0.15 hospital days per patient treated is likely to be similarly beneficial in other settings. Fluvoxamine is cost saving in locations where total hospital costs are >$738. Molnupiravir had an additional cost to the healthcare system of $404 per patient treated.

**Conclusions:** Fluvoxamine is cost-saving for COVID-19 outpatient therapy.

**Funding:** FastGrants and Rainwater Charitable Foundation

## Introduction

Recent evidence supports the use of oral fluvoxamine for the early, out-patient, treatment of symptomatic COVID-19. Two trials have reported on the efficacy of fluvoxamine in this population in reducing the need for hospitalization and/or progression to severe disease.[1, 2] These are currently the only published findings demonstrating treatment efficacy for oral medications for COVID-19. Repurposing existing medicines that are widely available as generic formulations and with well understood safety profiles, has particular appeal in supporting inexpensive scale-up during the pandemic.[3]

Fluvoxamine is a selective serotonin reuptake inhibitor (SSRI) and a Sigma-1 receptor (S1R) agonist.[4] There are several potential mechanisms for fluvoxamine in treatment of COVID-19 illness, including anti-inflammatory and possible antiviral effects.[5] Two published randomized clinical trials (RCT), one in the United States (n=150) and one from Brazil (n=1498), have indicated treatment effects on time to recovery as well as reduced emergency setting attendance and hospitalizations.[1, 2] Real world data also support the benefit of fluvoxamine.[6]

We applied a decision-analytic model to evaluate the most cost-effective strategy for out-patient treatment of adult patients presenting with symptomatic COVID-19 and known risk factors for disease progression. The paucity of options with which to inform other comparisons limits the analysis to two treatment strategies: (1) treatment with a standard 10-day course of fluvoxamine added to standard of care; or (2) current standard of care. As many as 20% patients at high-risk of disease progression require medical admissions, and that 32% of such admissions require intensive care unit (ICU) admission. [7] Invasive mechanical ventilation is required for 20% of ICU admissions.[8] Previous analyses report that, in the United States an uncomplicated hospitalization costs US$9,763, while a hospitalization with complications or a co-morbidity costs US$13,767.[8] Major complications or a co-morbidity increase estimated costs to US$20,292.[8] Furthermore, hospitalizations requiring the use of a ventilator are longer and more expensive, with the cost per admission requiring ventilator support for >96 hours topping US$88,000.[8] Given the magnitude of these costs, the results of a formal economic analysis of fluvoxamine in the early treatment of COVID-19 is a priority as there are negligible acquisition costs and safety is not a major concern. Herein, we report a cost-consequence analysis based on a meta-analysis of the available evidence from therapeutic trials.

## Methods

### Meta-analysis

We first conducted a meta-analysis of the three randomized clinical trials. We contacted the authors of each study and obtained trial results reporting on the number randomized, the number with medical admissions including hospitalization and ICU use, and the adverse event profiles. Two of the trials are published,[1, 2] and a third exists in preprint format (Clinicaltrials.gov number: NCT04668950). We applied a fixed-effects meta-analysis to inform the decision-model. We used established methods for interpreting meta-analysis including sensitivity analysis, heterogeneity assessment, and trial sequential analysis.[9]

### Decision-model

We applied the trial findings to the US hospital setting. Our modelled population included adults with confirmed COVID-19 infection who were at increased risk of progression to severe disease or hospitalization, based on established risk factors including age, obesity, and co-morbidities. The patient cohort within the model transitioned through the care pathway over the course of 28 days, as this was the duration of follow-up for the trials’ primary endpoints. The model reported the level of healthcare utilization by cohort, including extended emergency department use, hospital admission, ICU admission, total length of hospital stay, and discharge. Long-term consequences of COVID-19, i.e., with a clinical course exceeding 28 days, were not considered.

We applied a US healthcare system perspective in which third-parties (insurers) reimburse for healthcare services through bundled payments. Consequently, only direct medical costs, including but not limited to costs related to treatment acquisition, administration, and condition-related care, were considered. Productivity effects and other indirect costs were not relevant to this chosen perspective.

As the value of the primary endpoint of preventing disease progression was well established, we considered quality of life measures unnecessary. We assumed that avoidance of progression to severe disease is itself associated with increased quality of life and, considering the safety and tolerability of fluvoxamine,[2] that the results of a cost-utility analysis would show fluvoxamine is dominant, i.e. delivering greater health benefits at a lower total cost. Thus, a cost-consequence analysis was sufficient to address the study objective, with length of stay as an outcome of primary relevance to resource planning decisions in the US, but also interpretable for decision-making in other settings. Threshold analyses of the cost of hospital admission and ICU admission were conducted to identify the values at which the use of fluvoxamine became cost-neutral.

Given the limited time horizon of the trial data, and the fundamentally financial objectives of the analysis, we applied a decision-tree model, and a simple arithmetic model was constructed in MS Excel using the TreePlan add-in,(TreePlan Software, San Francisco, CA). The 28-day time horizon obviated the need for discounting of costs or effects. A schematic of the model is shown in **Figure 1**.

**Figure 1:**
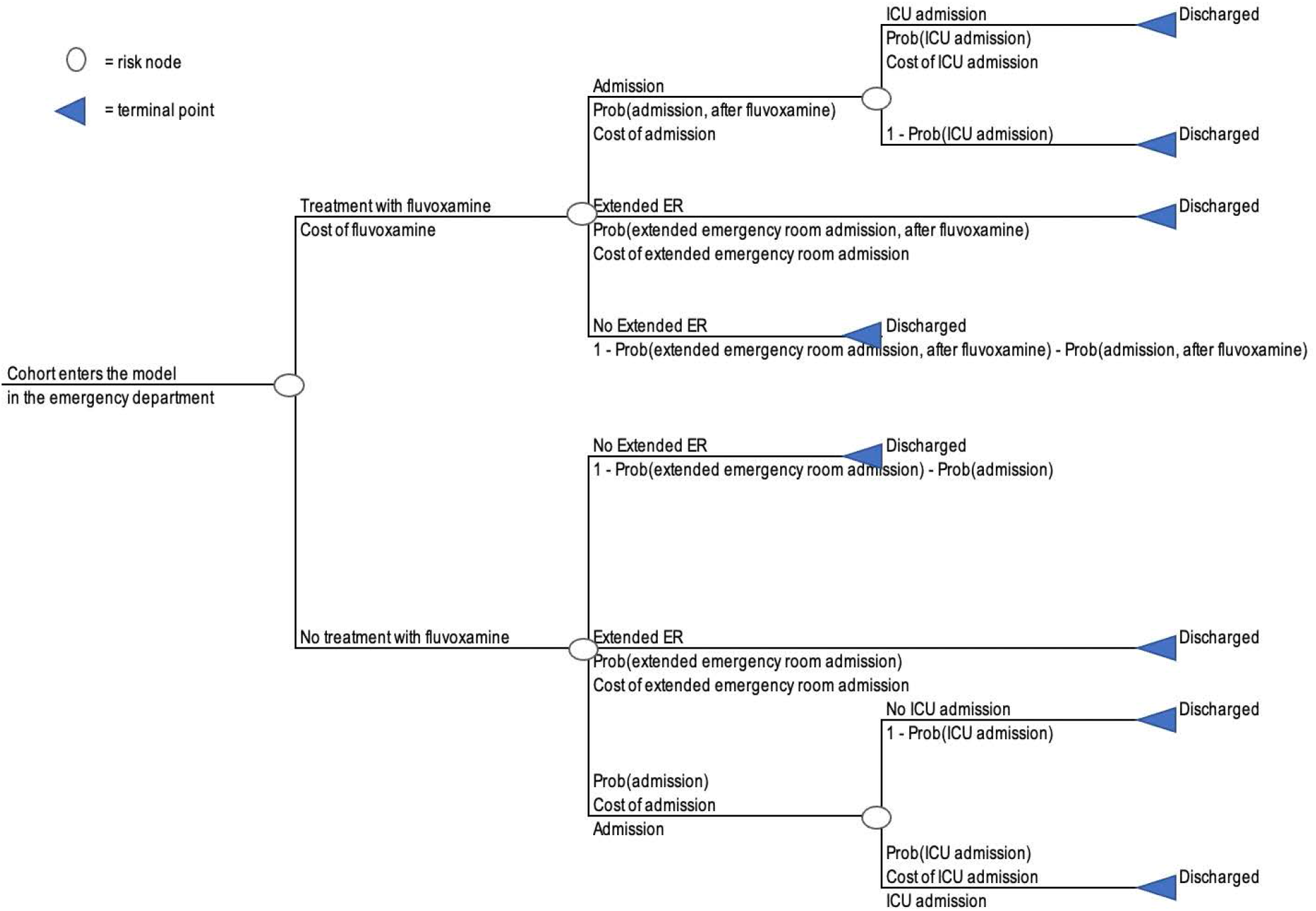
Model Schematic. Abbreviations: ER (Emergency Room); ICU (Intensive Care Unit)

### Input parameters

Our decision-tree model allowed for the calculation of expected costs for each of the treatment strategies, programmed as mutually exclusive sequences of events or pathways through which the patient cohort passes.[10, 11] Expected values were calculated by summing the pathway values, weighted according to the conditional probability of each sequence of events. Patient cohorts entered the model in the emergency department, with care escalating according to the rates reported in the intent-to-treat population of the trial publications, either in the form of extended emergency department stay or hospital admission. Since both cohorts entered the model in the out-patient setting, the costs of a routine ER visit were omitted, as they would be the same for both cohorts. The cost of the extended emergency visit is assumed to be 33% greater than the US$608.46 ER visit cost parameter in univariate sensitivity analysis. Patients who were at further risk of admission to the intensive care unit (ICU) would necessitate mechanical ventilation in a certain fraction of cases. The total length of stay is calculated for each strategy and reported with the cost results. Once hospitalization has occurred, the use of fluvoxamine is not expected to affect the likelihood of ICU admission, and so the same risk was applied to both model arms.

The clinical and economic data employed in the model are reported in Table 1. All costs were inflated to 2021 US dollars, using the Medical Care Inflation Calculator.[13] The age distribution of hospitalizations for COVID-19 and the probability of ICU for each of those age groups were obtained. [14] These values allowed the calculation of a weighted average for the probability of ICU admission, given hospitalization, shown in the clinical inputs table, and the weighted average cost for an ER visit; this value is then used to calculate the incremental cost associated with the extended ER visit.

**Table 1.**
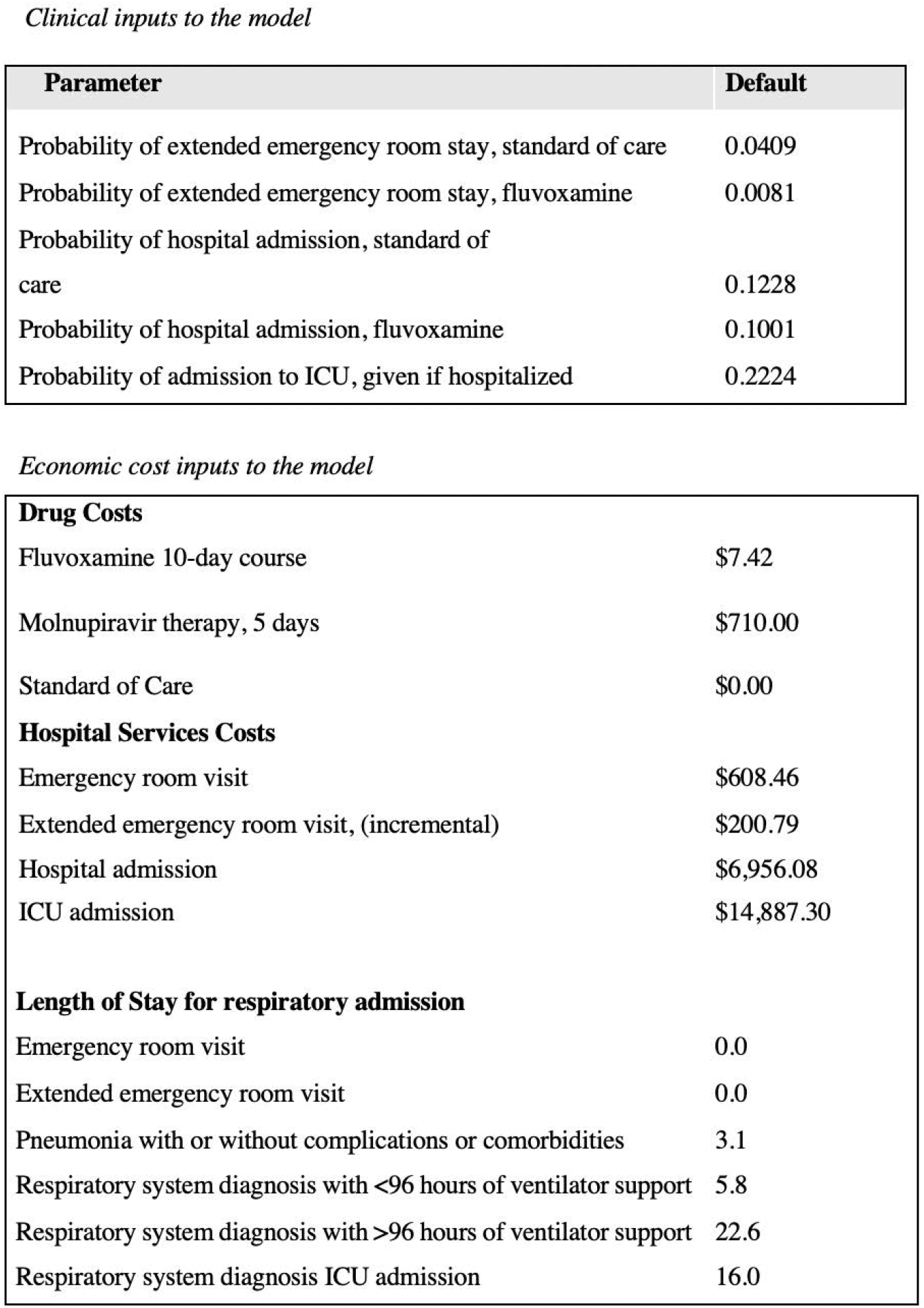
Clinical, economic and resource inputs to model, all 2021 USD.

Clinical inputs to the model
| Parameter | Default |
| --- | --- |
| Probability of extended emergency room stay, standard of care | 0.0409 |
| Probability of extended emergency room stay, fluvoxamine | 0.0081 |
| Probability of hospital admission, standard of care | 0.1228 |
| Probability of hospital admission, fluvoxamine | 0.1001 |
| Probability of admission to ICU, given if hospitalized | 0.2224 |

**Table 1.** Clinical, economic and resource inputs to model, all 2021 USD.
|  |  |
| --- | --- |
| **Drug Costs** |  |
| Fluvoxamine 10-day course | \$7.42 |
| Molnupiravir therapy, 5 days | \$710.00 |
| Standard of Care | \$0.00 |
| **Hospital Services Costs** |  |
| Emergency room visit | \$608.46 |
| Extended emergency room visit, (incremental) | \$200.79 |
| Hospital admission | \$6,956.08 |
| ICU admission | \$14,887.30 |
| **Length of Stay for respiratory admission** |  |
| Emergency room visit | 0.0 |
| Extended emergency room visit | 0.0 |
| Pneumonia with or without complications or comorbidities | 3.1 |
| Respiratory system diagnosis with <96 hours of ventilator support | 5.8 |
| Respiratory system diagnosis with >96 hours of ventilator support | 22.6 |
| Respiratory system diagnosis ICU admission | 16.0 |

Length of stay for each of the possible admission types were obtained from Rae et al.[8] Emergency visits, whether of normal duration or extended, were not assigned a hospitalization value. It has been estimated that 61% of patients admitted to ICU require ventilator support,[14] wherein we calculated a weighted average of 16 days. In addition to the primary assumptions reported in **Table 1**, one-way sensitivity analyses were conducted in order to assess the effect of individual parameter values and were presented in tabular form (**Table 2**) and as a tornado diagram (**Figure 2**). Inadequate data were available with which to inform a comparison of the efficacy of fluvoxamine and molnupiravir or the other therapies included in the Institute for Clinical and Economic Review’s Scoping Document for their Special Assessment of Outpatient Treatments of COVID-19.[7] As the competing direct-acting antivirals target earlier treatment of outpatients and are not yet provided in any published form, an indirect treatment comparison was not possible. However, in the interest of completeness,, a preliminary scenario analysis has been conducted to inform the potential cost-savings of molnupiravir, assuming that its initial efficacy results are borne out and lead to regulatory approval. A future report comparing all treatments will be pursued once sufficient data are available.

**Table 2.** One-way sensitivity analysis, inputs and results for Fluvoxamine

| Parameter | Base Case<br>Values | Model Inputs |  |
| --- | --- | --- | --- |
|  |  | Low | High |
| Probability (extended ER stay, fluvoxamine) | 0.0081 | 0.0042 | 0.0409 |
| Probability (Hospitalization, fluvoxamine) | 0.1001 | 0.08160 | 0.1228 |
| Extended ER cost | \$200.79 | \$100.40 | \$401.58 |
| ICU admission cost | \$14,887.30 | \$10,557.47 | \$21,943.28 |
| Fluvoxamine acquisition cost | \$7.42 | \$4.64 | \$12.95 |
| Hospital Admission cost | \$6,956.08 | \$5,564.87 | \$11,708.97 |

**Figure 2.**
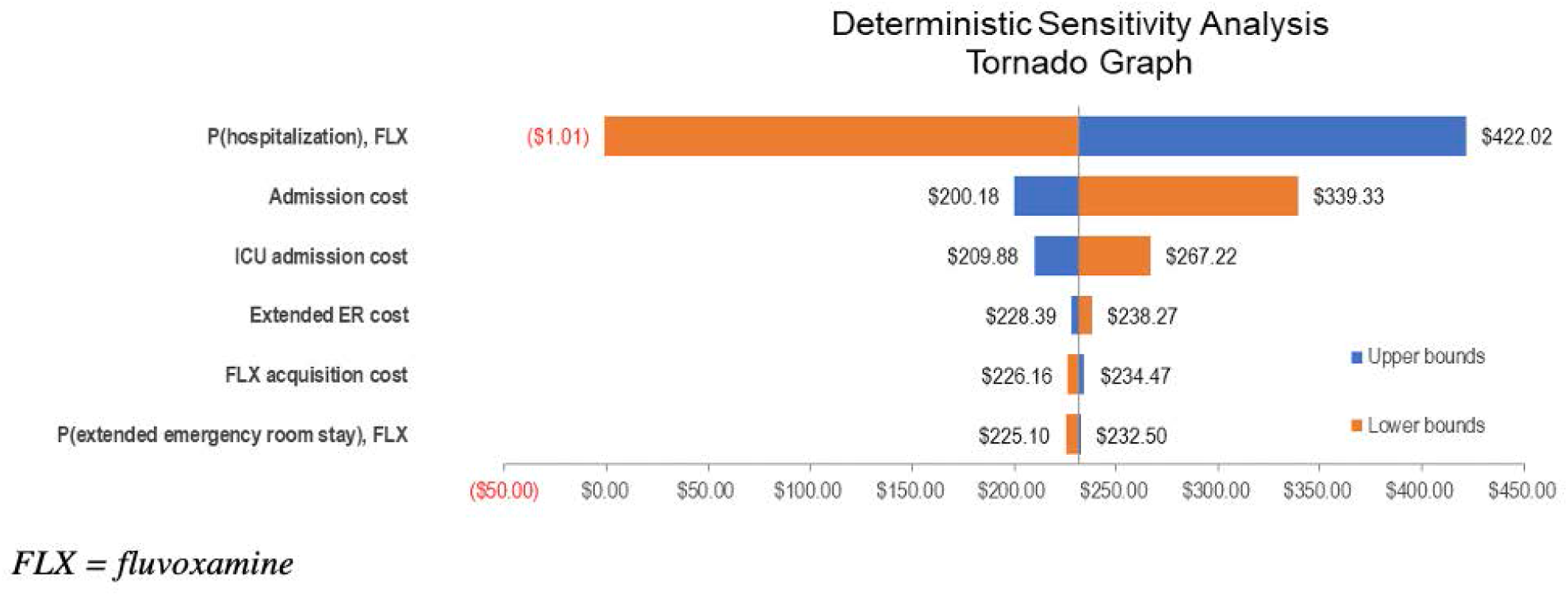
Tornado diagram for one-way sensitivity analyses

A series of one-way sensitivity analyses were conducted on the base-case analysis, as shown in the table below. The purpose of these analyses was to assess the effect of individual parameters on the results, using plausible alternate values for each. For the probability of an extended emergency visit with fluvoxamine, we used a rate reported in the meta-analysis as the lower bound, and the SOC rate for the upper bound. For the extended emergency visit cost, we used half and double the default value as the lower and upper bounds. For ICU admission cost, the lower bound used the reported value for pneumonia admission with no complications or comorbidities, while the upper bound used the same reported value for admission with major complication.13 The fluvoxamine acquisition cost is tested using the lowest and highest retail prices found for 100 units through online searches.[15] Finally, the cost of hospital admission was varied from that of 80% of the default value to the value calculated for severe pneumonia, weighted according to the published age profile of hospital admissions (2020).[14]

As molnupiravir is of considerable interest to formulary decision makers, we conducted a scenario analysis using the data available as of November 30, 2021. Pricing information was not yet available but has been reported to be approximately $710 per 10-day course in high-income countries.

Reporting of this analysis adheres to the Consolidated Health Economic Evaluation Reporting Standards (CHEERS) statement.[9]

## Results

The pooled estimate for hospitalization as defined in the primary trials was a relative risk of 0.74, 95% CI 0.56-0.98, I2=0% compared with placebo. The analysis is dominated by the TOGETHER trial, representing 90% of the weighting in the analysis. A sensitivity analysis examining only hospitalizations defined as tertiary hospitals (excluding emergency hospitals) indicated a pooled relative risk of 0.68 (95% CI, 0.52-0.83).

The primary results of our decision analysis are presented in **Table 3**, reflecting substantial cost-savings and reductions in hospital resource requirements associated with the use of fluvoxamine in the target population. The results suggest that the use of fluvoxamine will be cost saving by reducing length of stay by 0.15 days per person across the entire studied population. However, these results likely understate the true value of the intervention, as ICU admissions generally entail substantial additional costs in the year following after hospitalization.[14] Follow-up costs from admissions for acute respiratory distress syndrome, diagnosed in a large fraction of ICU-admitted patients, amount to an estimated US$28,133 in the year post-hospitalization, and sepsis, another common cause of ICU admission, incurs US$10,531 (2020 USD).[14] These costs were not captured in the 28-day time horizon. Additionally, extended ER visits are not assigned additional length of stay within the model, and incur only monetary costs, this may have the effect of underestimating the true resource savings and opportunities for efficiency presented by fluvoxamine use.

**Table 3:** Results of primary analysis. Abbreviations: SOC (standard of care)

| Cost Category | Fluvoxamine | SOC | Savings |
| --- | --- | --- | --- |
| Emergency Room | \$1.63 | \$8.22 | \$6.59 |
| Hospitalization | \$696.55 | \$854.09 | \$157.54 |
| Drug Costs | \$7.42 | \$0.00 | (\$7.42) |
| ICU Utilization | \$331.53 | \$406.51 | \$74.98 |
| Total Costs | \$1,037.13 | \$1,268.82 | \$231.69 |
| **Resource Use** |  |  |  |
| Regular admission days | 0.31 | 0.38 | 0.07 |
| ICU days | 0.36 | 0.44 | 0.08 |
| Total Hospital Days | 0.67 | 0.82 | 0.15 |

As is evident from the tornado diagram (**Figure 2**), the cost of hospital admission is the key driver of the result, as this is both a substantially expensive and frequent event, reported in approximately 15% of studied patients in both the TOGETHER trial,[2] and 10% in the recently-reported molnupiravir trial.

In order to inform resource allocation decisions in other settings, we conducted two threshold analyses, for the cost of hospital admission and the cost of ICU admission, in order to determine the values at which the use of fluvoxamine would be expected to be cost-neutral. As the values were tested independently, there were no positive values for either parameter that would make fluvoxamine use cost-neutral. However, setting the value for extended ER visit to zero yielded a threshold value of $328 for hospital or ICU admissions. Alternatively stated, fluvoxamine would be cost saving wherever the total cost of COVID-19 hospitalization exceeded $328.

An important issue relating to the external validity of the TOGETHER trial is the unvaccinated status of its subjects. It has been reported that hospitalization is several times lower in fully vaccinated people,[16] so it is of interest to determine whether fluvoxamine would remain cost-saving if hospitalizations were, e.g., one-third less frequent. By reducing the likelihood of both extended ER visits and hospitalizations by one third for each arm, thus maintaining the same relative effect, the model suggests that fluvoxamine remains cost-saving, in the amount of $72.28, reducing hospitalization days by 0.05. As molnupiravir is of considerable interest to formulary decision makers, we conducted a scenario analysis using the data available as of November 30, 2021.[17] Pricing information was not yet available, but has been reported to be approximately $710 per 10-day course in wealthier nations.[18] The primary outcome of the molnupiravir trial differs from that of the TOGETHER trial, being a composite of hospitalization or death, and its efficacy estimates have been revised downwards since initial announcements. The approximate 30% risk reduction for the composite outcome of hospitalization or prolonged ER visits for molnupiravir is of similar magnitude as that of fluvoxamine but at approximately 100 times the cost.[17]

At the anticipated price of $710 per 5-day course of molnupiravir treatment, assuming that the molnupiravir effect on extended ER visits is the same as fluvoxamine (not publicly reported), and based on its most recent efficacy estimates, molnupiravir is not cost-saving at an additional cost of $404 per high-risk patient treated (Table 4). Importantly, low-income countries expected to be cost-saving, despite saving more hospitalization time than fluvoxamine. This may be able to access molnupiravir at a greatly reduced price, attributable to the combination of higher cost and the lower overall event rate reported in the molnupiravir trial. An additional caveat regarding molnupiravir’s anticipated clinical utility is warranted, as there appear to be important safety signals associated with molnupiravir which may limit its uptake in men and women of child-bearing potential.[17]

**Table 4:**
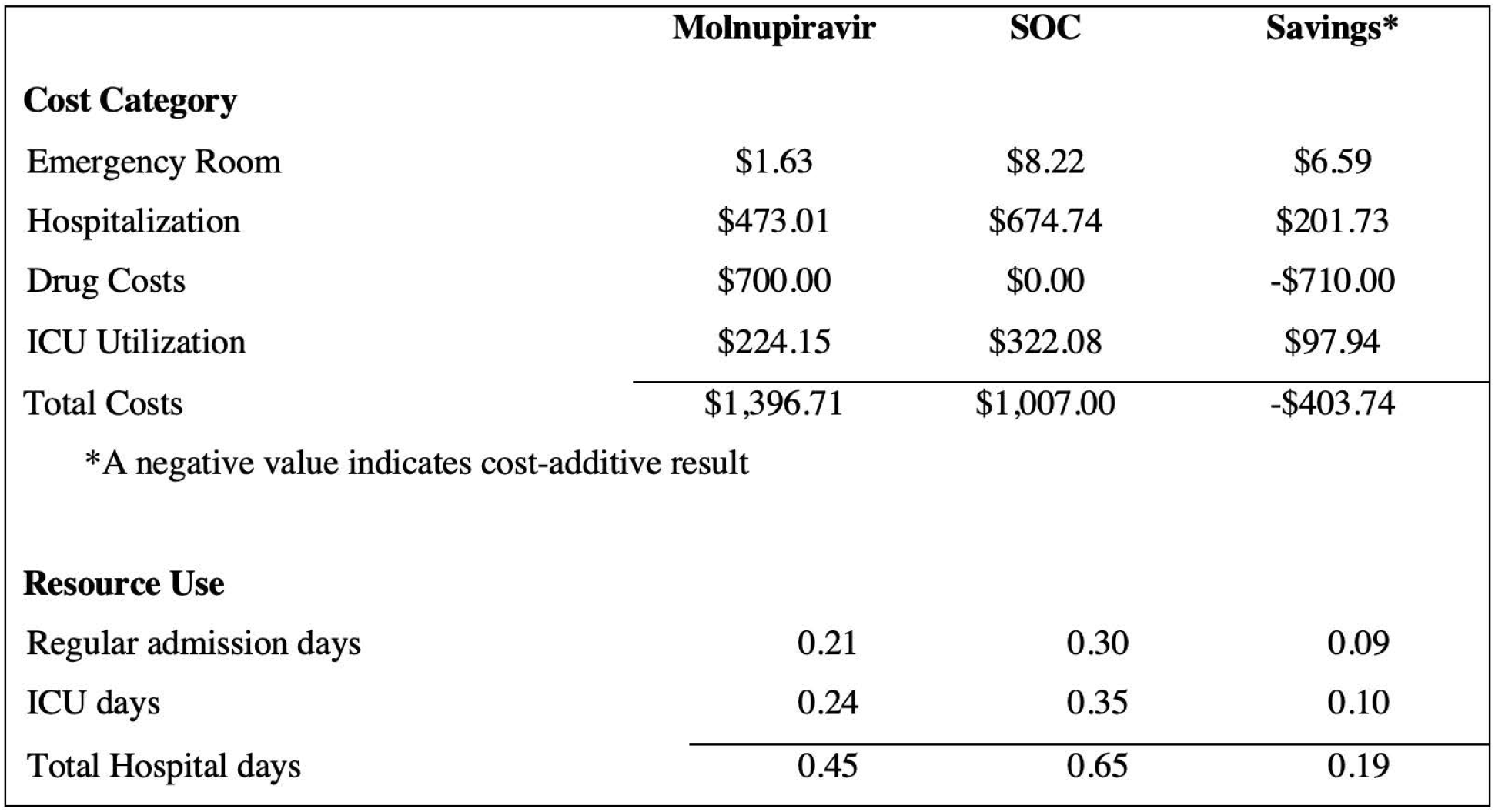
Results of primary analysis. Abbreviations: SOC (standard of care)

|  | Molnupiravir | SOC | Savings* |
| --- | --- | --- | --- |
| **Cost Category** |  |  |  |
| Emergency Room | \$1.63 | \$8.22 | \$6.59 |
| Hospitalization | \$473.01 | \$674.74 | \$201.73 |
| Drug Costs | \$700.00 | \$0.00 | -\$710.00 |
| ICU Utilization | \$224.15 | \$322.08 | \$97.94 |
| Total Costs | \$1,396.71 | \$1,007.00 | -\$403.74 |
| *A negative value indicates cost-additive result |  |  |  |
| **Resource Use** |  |  |  |
| Regular admission days | 0.21 | 0.30 | 0.09 |
| ICU days | 0.24 | 0.35 | 0.10 |
| Total Hospital days | 0.45 | 0.65 | 0.19 |

## Discussion

This study found that administration of a 10-day course of fluvoxamine to symptomatic, high-risk COVID-19 outpatients in the emergency room is substantially cost-saving in the US. The cost-savings expected from fluvoxamine adoption are unsurprising, given the reduction in incidence of hospitalization expected from the administration of a comparatively inexpensive, generic medication. Whether fluvoxamine becomes a standard of care will differ across settings and current findings, together with the results of further clinical trials will inform global guidance.

There are strengths and limitations to our analysis. Strengths include that we obtained results from each of the trials to inform both the meta-analysis and the decision-analysis. As with all trials, the demographic and disease characteristics of the population enrolled may differ from those seen in other health systems, particularly in vaccination status. However, subgroup analyses uniformly demonstrate favorable results for fluvoxamine.[2] This suggests that heterogeneity of patient characteristics is unlikely to undermine the validity of the conclusions of this analysis. Nonetheless, two sources of uncertainty affect the selection of appropriate patients. First, the TOGETHER study, which dominated the meta-analysis, was conducted among predominantly unvaccinated patients, and further evidence is therefore needed to inform the clinical value of fluvoxamine among vaccinated populations. Among vaccinated persons with breakthrough infections aged >50 years, they may have 2-3.5 fold lower risk of hospitalization than unvaccinated populations.[16] Even with this reduction in risk of severe disease, fluvoxamine would be cost beneficial. However, a naïve scenario analysis based on reduced treatment escalation rates suggested that the primary findings remain robust. Second, enrolled patients were not already receiving treatment with fluvoxamine or medications within the SSRI class. Further research is therefore needed to determine whether patients already receiving such therapies should be expected to experience the same benefits observed in TOGETHER. The model accomplishes its objective without accounting for differential health outcomes, as might be measured by “equal-value life years gained”, or quality-adjusted life years gained. However, the inclusion of such outcomes in a future model would likely show fluvoxamine as dominant, i.e., producing more population health at a lower overall cost. The current model does not permit consideration of a wider effect on the health system’s capacity, or healthcare personnel, but it is likely that the avoidance of disease progression and attendant resource consumption would have substantial benefits in these areas, and clearly this assessment is needed. In conclusion, by reducing the total costs and reducing the need for escalation of care, the use of fluvoxamine in emergency departments is substantially cost-saving in the treatment of symptomatic COVID-19 patients with known risk factors. Given fluvoxamine’s tolerability, ease of use, affordability and easy access, this finding has the potential to positively influence the health system responses to the clinical management of COVID-19, which in turn can influence the balance of risks and benefits for adopting broader COVID-19 containment measures.

## Data Availability

Data used in this paper will be made available following publication of this manuscript to interested investigators through the International COVID-19 Data Alliance after accreditation and approval by the authors.

## Acknowledgments

We acknowledge our partners and funders in the preparation of this publication.

